# Generation of antibodies against COVID-19 virus for development of diagnostic tools

**DOI:** 10.1101/2020.02.20.20025999

**Authors:** Maohua Li, Ronghua Jin, Ya Peng, Cuiyan Wang, Wenlin Ren, Fudong Lv, Sitao Gong, Feng Fang, Qianyun Wang, Jianli Li, Tong Shen, Hunter Sun, Lei Zhou, Yali Cui, Hao Song, Le Sun

## Abstract

The COVID-19 China coronavirus started in Dec 2019 was challenged by the lack of accurate serological diagnostic tool for this deadly disease to quickly identify and isolate the infected patients. The generation of COVID-19-specific antibodies is essential for such tasks. Here we report that polyclonal and monoclonal antibodies were generated by immunizing animals with synthetic peptides corresponding to different areas of Nucleoprotein (N) of COVID-19. The specificities of the COVID-19 antibodies were assessed by Western Blot analysis against NPs from COVID-19, MERS and SARS. Antibodies were used for immunohistochemistry staining of the tissue sections from COVID-19 infected patient, as a potential diagnostic tool. A Sandwich ELISA kit was quickly assembled for quantitation of the virus/NP of COVID-19 concentrations in the vaccine preparations. Development of POCT is also aggressively undergoing.

---

Recently, a novel coronavirus, named COVID-19, broke out from Wuhan, China, had spread rapidly around the country and now the rest of world. The pneumonia symptoms manifested by this virus are similar to that of the SARS coronavirus (SARS-CoV), which lead to acute respiratory syndrome. Until Feb 14^th^, 2020, 63,932 people confirmed to be infected by COVID-19 with 1,596 deaths in China^1^. Comparatively, SARS-CoV infected a total of more than 8,000 persons with 774 deaths in 37 additional countries in 2003^2^. More seriously, the infections among doctors and family clusters demonstrated that it could transmission among humans^3-6^, more aggressively than what were observed with the SARS outbreak in 2003. Making the issue even worse, the virus could be widely transmitted by asymptomatic virus-carriers to people in close contact^7^ and some of the patients will still carry the virus even after they recovered from the diseases. RT-PCR has been used since the first day to screen the suspected and now used for confirmation of diagnosis for COVID-19 infection due to the lack of immunologic diagnostic assays for the new virus^8^. Molecular diagnosis requires both special laboratory setting and very skilled laboratory personnel, which are both highly limited in Wuhan and the heavily hit surrounding epidemic areas. False negative rate was alarming at 30-40% with RT-PCR of throat swabs due to many different reasons. It is urgent to develop rapid, fast and simple screening tools to find the “moving source of virus” and quarantine them.

COVID-19 contains Nucleoprotein (N-protein), Spike glycoprotein (S-protein), Envelope protein (E-protein) and Membrane protein (M-protein)^9^. Ideally, developing antibody against the S protein will be the best approach since S plays very important roles in virus entering by binding to the target receptor on the cell surface. However, S protein is a heavily glycosylated protein^10^ and many groups have tried to express the COVID-19 S protein in mammalian cells with limited success. On the other hand, N-protein is produced at very high levels in the virus infected cells and is considered to be a good candidate for clinic diagnosis^11^.

Based on our past success experiences of generating neutralizing monoclonal antibodies against EV71 virus and human DKK2 using synthetic peptides as immunogens^12,13^, we identified a few sequences in COVID-19’s NP that could be linear epitopes with promising antigenicity on the outer surfaces of NP. In this report, we present that, in events like this outbreak of COVID-19 virus, using synthetic peptides as immunogens may offer us a way to develop serological diagnostic tools in a timely manner. Our work also demonstrated that by working closely with scientists, clinicians, biotechnology contract research organization (CRO), IVD manufactures companies, NGOs and many different government agencies, together we could quickly translate the genomic information of emerging pathogens to many useful research and diagnostic tools to combat the deadly diseases such as the on-going global crisis of OCVID-19 virus.

## MATERIALS AND METHODS

### Peptides synthesis and antibody generation

Peptides were synthesized at DentriPro Bioscientific Ltd. Each peptide was chemically linked to the carrier protein mcKLH through a sulfide-linker.

Balb/c mice and New Zealand White rabbits were immunized with KLH-conjugated synthetic peptides. Bleeds were tested for titers against unconjugated peptides and purified NP by ELISA.

Splenocytes from the immunized mice were fused with the mouse myeloma cell line SP2/0 and cultural supernatant from individual hybridoma clones were screened against NP by ELISA. Bulk mAbs were produced using stationary bioreactors. The concentrations of purified IgG were determined by their absorbance at OD_280_.

### Enzyme-linked immunoassay and Western Blot analysis

Wells were coated with 1μg/mL of un-conjugated peptides or recombinant protein, blocked with 5% skim-milk/PBS, and incubated with bleeds or culture supernatants, then probed with HRP-conjugated secondary antibodies. After final washes, HRP substrate TMB solution was added and absorbencies were determined at 450nm with a microplate reader.

Recombinant NPs from COVID-19, SARS and MERS were provided by Chinese CDC and Inst. of Microbiology, Chinese Academy of Science. Proteins were electrophoresed using 12% SDS-PAGE. Proteins were transferred onto nitrocellulose membranes and blocked with 5% non-fat milk for 2 h, then stained with primary antibodies for 1h, then detected with HRP-conjugated secondary antibodies for 50 min. After final wash, the blots were analyzed using SuperSignal West Pico Chemiluminescent Substrate (Life Technologies).

### Immunohistochemistry Staining

Following Novolink Polymer’s instruction, dewaxed lung tissue sections were immersed in 0.3% H_2_O_2_ to reduce the background of endogenous peroxidase, incubated with the primary antibodies for 60 min at 37 °C, treated with enhancer for 30 min at 37 °C, then probed with HRP-conjugated secondary antibody for 30 mins at 37 °C. The DAB substrate was used to develop the color, followed by counter-staining and dehydrating the sections. The sections were examined under microscope.

### Sandwich ELISA and Rapid Immunochromatographic Assay(RICA)

Anti-COVID-19 NP rabbit pAbs were conjugated with biotin according to the manufacturer’s instructions.

For Sandwcih ELISA, as described previously^14,15^, each well was coated with different rabbit anti-COVID-19 NP pAbs and blocked with 3% BSA in PBS. Samples were diluted in 3% BSA-PBS and incubated in the wells for 1 h, probed with the Biotin-conjugated rabbit anti-COVID-19 NP pAbs diluted in 3% BSA-PBS, then detected with HRP-Streptavidin. After washes, chromogenic HRP substrate TMB was added and the reaction was stopped with 0.1 M H_2_SO_4_ and absorbance was measured at 450 nm.

For RICA, Anti-COVID-19 NP rabbit pAb (Pept-4) was dispensed on a nitrocellulose membrane as a “test” (T) line, and goat anti-rabbit IgG was fixed as a “control” (C) line. Quantum Dots (QD) pre-coated with a layer of silica were covalently conjugated with the Anti-COVID-19 NP rabbit pAb (Pept-3) and distributed on a glass fiber. The sample pad, conjugate pad, nitrocellulose membrane, and absorbent pad were attached to a lamination pad and cut to fit in a plastic cartridge. Sera were diluted with Tris buffer containing 1% NP-40 and two drops of the sample were loaded to the test strip and wait for 15 minutes then scanned with SkanFlexi X200 with the associated software to calculate the concentration of the antigen based on the intensities the T and C lines.

## RESULTS

### Design and synthesis of peptides as immunogens for generation of COVID-19-specifc antibodies

NPs of coronavirus are shared with very high homology between all seven coronaviruses. Cross-reactivity among NPs of human coronaviruses were reported by Yu’s group^16^. In this study, 5 linear sequences, SDNGPQSNQRNAPRITFG (Pept-1,a.a.2-19), PSDSTGSNQNGERSGARSKQ (Pept-2, a.a.20-39), PGSSRGTSPARMAGNGGDAALA (Pept-3, a.a.199-220), PKKDKKKKADETQALPQRQKK (Pept-4, a.a.368-388) and GGSADSTQA (Pept-5, a.a.413-419) were selected as the potential linear epitopes on the outer surfaces of COVID-19 NP.

Normally, the rotations/conformations of the very ends of proteins are more close to the ones of the synthetic peptides so the antibodies generated with synthetic peptides corresponding to the NT and CT of the target protein have higher possibilities to bind to the native target proteins. Pept-1 (SDNGPQSNQRNAPRITFG) is located at the very end of N terminal and Pept-5

(GGSADSTQA, a.a.413-419) are the last 9 amino acids of COVID-19 NP. Pept-1 shares 88.9% identify with the SARS NP, and no homology to the MERS’ and the other 4 coronaviruses’ NP. Pept-5 is 100% identical in COVID-19 and SARS, no homology with the rest 5 viruses. The other three sequences were selected based more on the specificity and immunogenicity other than the locations in the protein. Pept-2 has only 70% identity to SARS’s in amino acid composition but are not in continued sequence and the antibodies generated by Pept-2 will have the highest possibility to be COVID-19 specific. Pept-3 has 77% homology with SARS and 50% identity to MERS at the very end of NT of the peptide. Pept-4 has the strongest immunogenicity and its NT has very high homology with both SARS and MERS.

### Generations of pAbs and mAbs to COVID-19 NP

Five groups of mice, 3 mice per group, were immunized with the 5 KLH-peptides respectively. On day 12, the tail bleeds were collected and examined by ELISA against both the corresponding peptides and the full length COVID-19 NP. As shown in Table 1-1, the mice immunized with Pept-4-KLH not only generated strong immune responses against the immunizing peptide, but also developed antibodies bound to COVID-19 NP. On day 13, the spleenocytes were isolated from the immunized mice and fused with SP2/0 mouse myeloma cells to generate mouse hybridoma cells.

**Table 1-1.** Titers of the peptide immunized mice tail bleeds by ELISA

| Immunogen |  | 1/500 | 1/1000 | 1/5000 |  | 1/500 | 1/1000 | 1/5000 |
| --- | --- | --- | --- | --- | --- | --- | --- | --- |
| Pept-1-KLH | 1# | 0.048 | 0.020 | 0.010 | 2# | 0.189 | 0.096 | 0.022 |
|  | 3# | 0.030 | 0.014 | 0.010 |  |  |  |  |
| Pept-2-KLH | 1# | 0.120 | 0.077 | 0.016 | 2# | 0.009 | 0.011 | 0.007 |
|  | 3# | 0.009 | 0.011 | 0.007 |  |  |  |  |
| Pept-3-KLH | 1# | 0.034 | 0.021 | 0.010 | 2# | 0.081 | 0.059 | 0.012 |
|  | 3# | 0.019 | 0.013 | 0.009 |  |  |  |  |
| Pept-4-KLH | 1# | 0.943 | 0.472 | 0.112 | 2# | 0.607 | 0.323 | 0.067 |
|  | 3# | 1.146 | 0.656 | 0.120 |  |  |  |  |
| Pept-5-KLH | 1# | 0.011 | 0.007 | 0.006 | 2# | 0.015 | 0.009 | 0.010 |
|  | 3# | 0.024 | 0.015 | 0.008 | PC | 1.125 |  |  |

Notes: The mice tail bleed were collected and evaluated the titers against full length NP by ELISA.

Weak immune reactions against corresponding peptides were observed with mice immunized with Pept-1-KLH or Pept-3-KLH but no significant reactions with the full length COVID-19 protein were observed. Immunization with Pept-2-KLH and Pept-5-KLH failed to induce any significant immunization responses at such short time of period.

Started the same time, three groups of New Zealand White rabbits (two each) were immunized with Pept-2-KLH, Pept-3-KLH and Pept-4-KLH respectively. On day 21, sera were examined against the NP by ELISA. As shown in Table 1-2, similar to what been observed in mice, rabbits immunized with Pept-4-KLH generated the best immune responses against the COVID-19 NPs, while moderate antibody titers were also observed in rabbits immunized with Pept-2-KLH and Pept-3-KLH. All animals’ titers increased significantly from day 21 to day 28, but some reached plateau after day 28. Polyclonal antibodies were purified using Protein A column for preparation of conjugations and pairing.

**Table 1-2.**
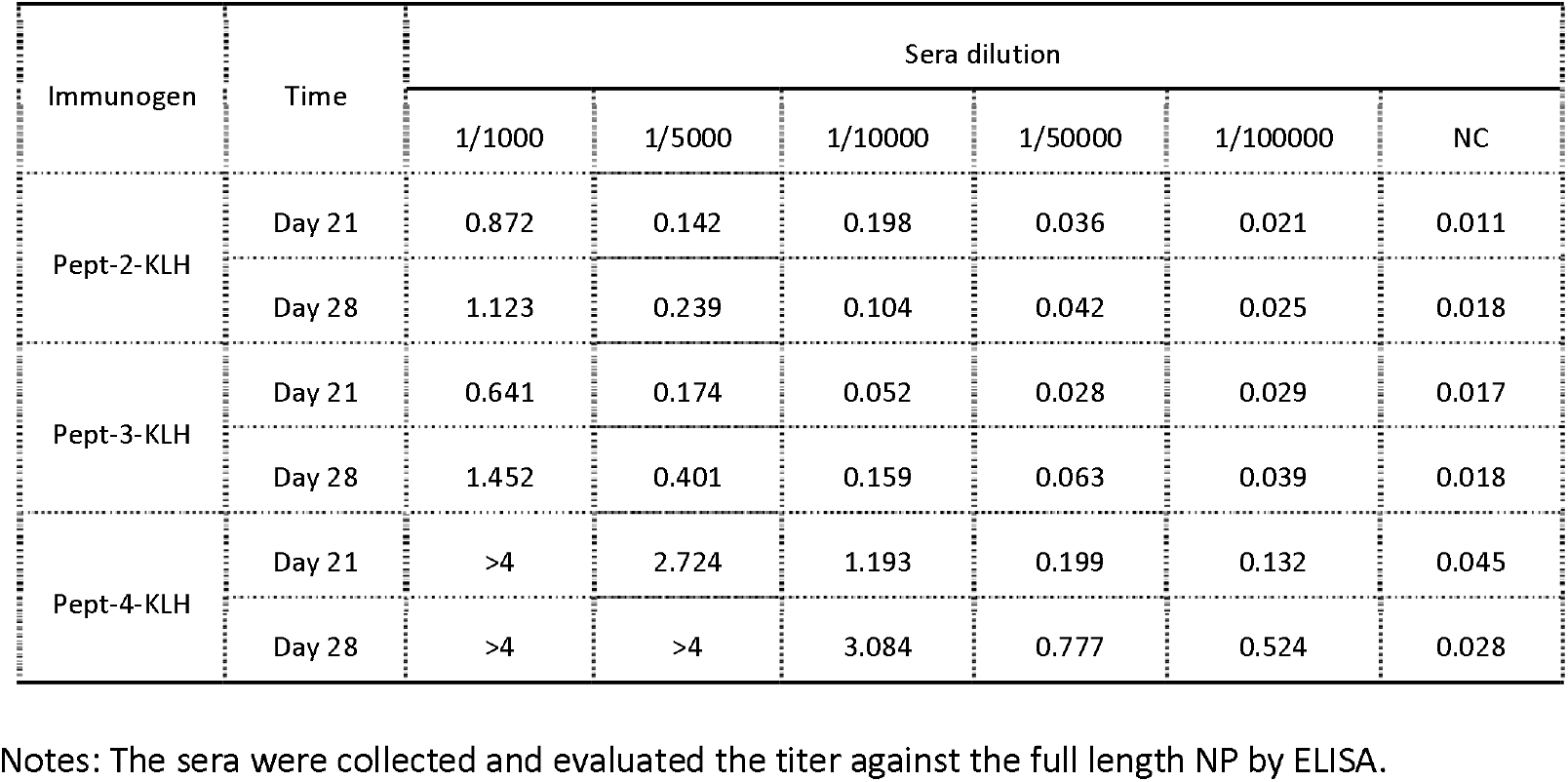
Titers of the rabbit sera against NP

| Immunogen | Time | Sera dilution |  |  |  |  |  |
| --- | --- | --- | --- | --- | --- | --- | --- |
|  |  | 1/1000 | 1/5000 | 1/10000 | 1/50000 | 1/100000 | NC |
| Pept-2-KLH | Day 21 | 0.872 | 0.142 | 0.198 | 0.036 | 0.021 | 0.011 |
|  | Day 28 | 1.123 | 0.239 | 0.104 | 0.042 | 0.025 | 0.018 |
| Pept-3-KLH | Day 21 | 0.641 | 0.174 | 0.052 | 0.028 | 0.029 | 0.017 |
|  | Day 28 | 1.452 | 0.401 | 0.159 | 0.063 | 0.039 | 0.018 |
| Pept-4-KLH | Day 21 | >4 | 2.724 | 1.193 | 0.199 | 0.132 | 0.045 |
|  | Day 28 | >4 | >4 | 3.084 | 0.777 | 0.524 | 0.028 |

Notes: The sera were collected and evaluated the titer against the full length NP by ELISA.

### Screening of mouse anti-COVID-19 monoclonal antibodies

On day 21, culture supernatants from 752 stable monoclonal hybridomas were screened against NPs, and more than 100 positive clones were obtained. On day 25, the culture supernatants not only were tested for the stable productions of monoclonal ant-COVID-19 NP antibodies but also examined their cross-reactivity towards the NPs from SARS and MERS (Partial of the data were shown in Table 1-3). As you can see, clone 6F10 mAb had the highest titer against COVID-19’s NP with minimal cross-reactivity to SARS’s NP, and no significant reactivity to MERS’s NP. On the contrary, clone 3A3 and 1F7 reacted equally well to NPs from COVID-19, SARS and MERS. Clone 3A10 and 8C3 recognized NPs from COVID-19 and SARS, but not the one from MERS. Our data suggested that even with such short of sequence, there could be many different antigen epitopes that can generate different mAbs with different specificities.

**Table 1-3.** Cross-reactivity Test of the supernatants from different clones by ELISA

| Clone | 1E10 | 1F7 | 3A1 | 3A10 | 3A1 | 3A10 | 3A11 | 3A3 | 4C4 | 6F10 | 6H1 | 7A1 | 7H1 | 8C3 |
| --- | --- | --- | --- | --- | --- | --- | --- | --- | --- | --- | --- | --- | --- | --- |
| COVID-19 | 0.385 | 0.223 | 0.335 | 0.309 | 0.335 | 0.309 | 0.449 | 0.305 | 0.205 | 0.803 | 0.372 | 0.335 | 0.387 | 0.396 |
| SARS | 0.240 | 0.269 | 0.442 | 0.372 | 0.442 | 0.374 | 0.158 | 0.515 | 0.090 | 0.101 | 0.097 | 0.018 | 0.209 | 0.214 |
| MERS | 0.087 | 0.246 | 0.137 | 0.096 | 0.137 | 0.096 | 0.008 | 0.310 | 0.183 | 0.050 | 0.124 | 0.115 | 0.081 | 0.035 |

Notes: The supernatants were collected and evaluated with NPs from three different strains of coronaviruses (COVID-19, SARS and MERS) by ELISA.

### Western Blot Analysis and Immunohistochemistry staining

As shown in Fig.2A and 2B, tested against purified recombinant NPs from COVID-19, SARS and MERS, both rabbit pAbs (Pept-3) and mAb 6F10 (Pept-4) reacted with COVID-19 NP very well with a clear band around 50 kDa while the rabbit pAbs showed some extend level of cross-reactivity with NPs from SARS and MERS (recombinant MERS was GST-tagged). Most importantly, mouse mAb 6F10 showed very good specificity to COVID-19 NP with minimal cross-reactivity to the ones from SARS and MERS.

**Figure 1.**
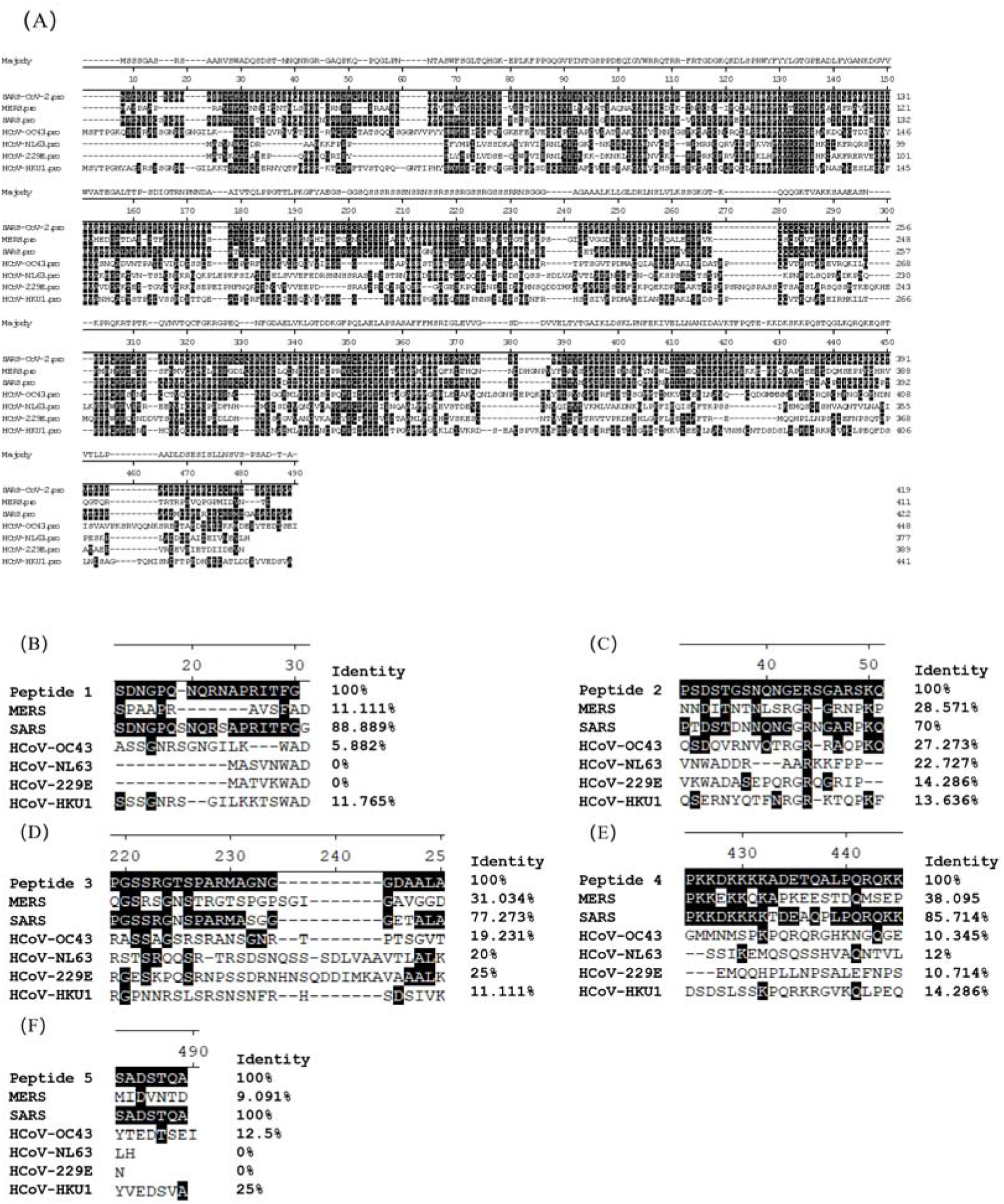
The alignment of peptides and full length NPs from COVID-19 and six other human infected coronaviruses. Alignments of full length NP (A), peptide 1(B), peptide 2(C), peptide 3(D), peptide 4(E) and peptide 5(F) from COVID-19-NP with NPs from other strains of coronaviruses. The sequences of NPs from different strains of coronaviruses were selected from NCBI. The protein sequences were aligned by clustal W method.

**Figure 2.**
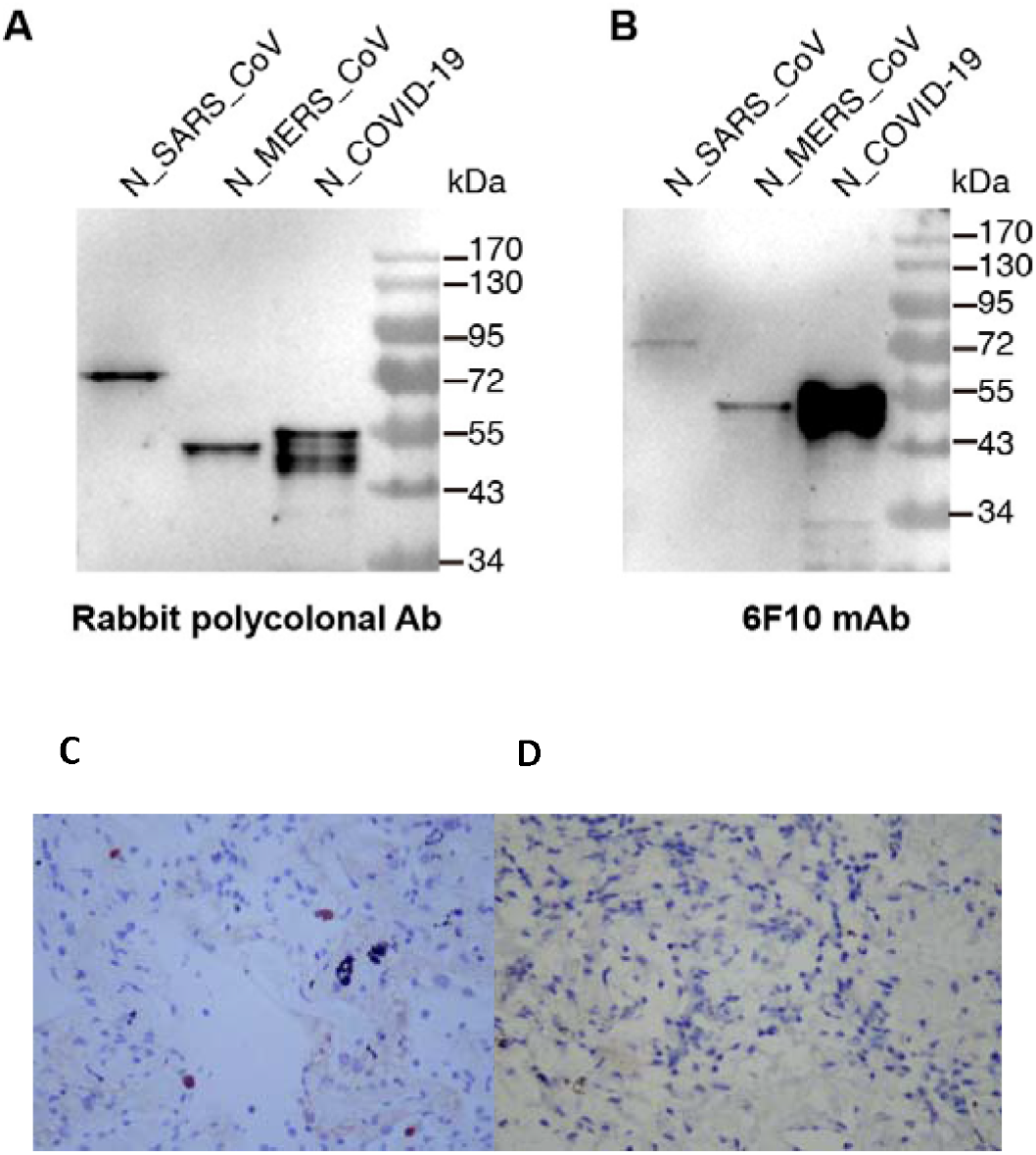
Detection of the NP with antibodies by different methods. Western blot against NP from different coronaviruses. Different NPs were immunoblotted with rabbit pAbs (Pept-3, A) or mAb 6F10 (B), then detected with HRP-conjugated secondary antibodies, and developed by Enhanced Chemiluminescent. And IHC staining of biopsy sections with anti-COVID-19 pAbs were evaluated. (C) The sample from the patients confirmed with SARS-CoV-2 by RT-PCR; (D) The sample from the patients diagnosed with non-specific lung infection. All the samples were stains with anti-COVID-19 pAbs (Pept-2) and detected with horseradish peroxidase-conjugated goat anti-rabbit IgG.

The biopsy samples of pulmonary sarcoidosis (after ablation) were collected from patients either confirmed with COVID-19 virus infection by RT-PCR or diagnosed with non-specific lung infection. The biopsy samples were stained with anti-COVID-19 pAbs (Pept-2). As shown in Fig. 2C and 2D, under the microscope view (400X), there were four cells with very distinguish dark-brown staining in the COVID-19 infected patient’s biopsy tissue. The staining was evenly distributed in the cytosol with inclusion body appearances. No nuclear staining. Those four stained cells were located inside the alveolar cavity, and look like macrophages. No brown staining was observed in the negative control sample from the patient with non-specific infection.

### Development of Sandwich ELISA and rapid immunochromatographic assays

The 96-well plate were coated with different NP pAbs, and paired with different Biotin-labeled NP pAbs. The best combination was coating with anti-COVID-19 NP pAbs (Pept-3) and paired with Biotinylated anti-COVID-19 NP pAbs (Pept-4).

A Sandwich ELISA kit was quickly assembled for quantitation of inactivated COVID-19 NP/virus concentration in vaccine. The assay kit had a limit of detection around 100ng/mL with a linear range between 200-1600ng/mL (see Fig. 3A).

**Figure 3.**
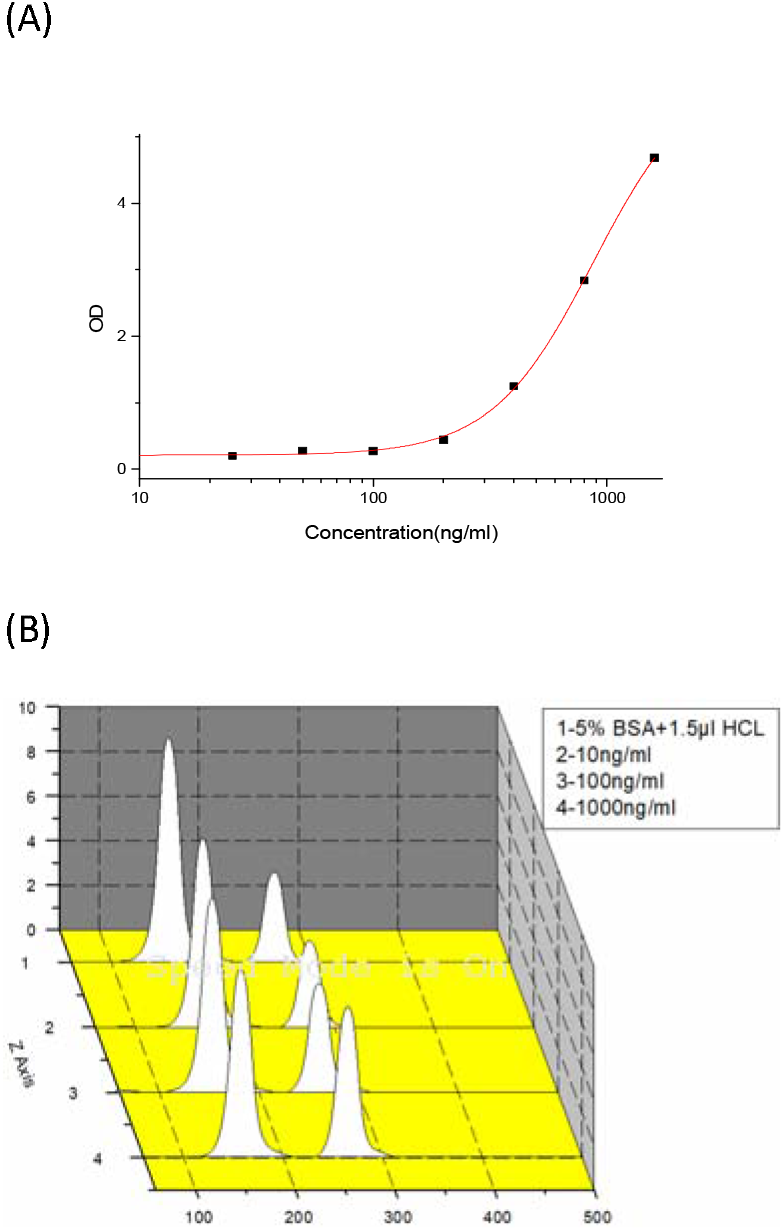
Development of Sandwich ELISA and rapid immunochromatographic assays. Detection limit of antigen-capture Sandwich ELISA. COVID-19 NP was used for examination the limit of detection, and the absorbance was measured at 450 nm with a plate reader; (B) Development of rapid immunochromatographic assays. The peaks of T and C measured using QD biosensor.

To race against the time, more than 6 different immunochromatographic technology platforms were attempted, including Quantum Dots-chromatographic assay, magnetic immuno-chromatographic assay (MICA). As shown in Fig. 3B, the limit of detection of QD-based ICT was about 10ng/mL. Further improvement of detection sensitivity from many different aspects has been aggressively pursuing.

## DISCUSSION

In this report, we demonstrated that, in event like this outbreak when only the gene sequence information was available, we could design and synthesis the peptides as the immunogens to quickly generate both polyclonal and monoclonal antibodies for development of diagnostic tools against the emerging diseases. Polyclonal anti-COVID-19 NP antibodies were successfully used as pathology tool for diagnosis of COVID-19 infection. The Sandwich ELISA kit has been used to help the vaccine developers to monitor the virus expression levels in the cultures in the process of optimization of manufacture parameters.

## Data Availability

The data used to support the findings of this study are available from the corresponding author upon request.

## AUTHOR CONTRIBUTIONS

### Author Contributions

ML, FF, CW, QYW, STG contributed the antibody generation and characterization. WLR, TS, HS contributed in the bioinformatics and peptide antigen design. JLL contributed in the Sandwich ELISA. YP and HS contributed the recombinant NP production and WB analysis. RHJ and FDL contributed the immunohistochemistry study. LZ and YLC contributed the ICT development.

## Acknowledgments

This work is supported by Research Grants from Beijing Science and Technology Commission, and Bill & Melinda Gates Foundation to Le Sun. This work was also supported by the National Natural Science Foundation of China (NSFC) (81702015), and the National Science and Technology Major Project (2018ZX10733403) to H.S. We thank Prof. Wenjie Tan from National Institute for Viral Disease Control and Prevention, Chinese Center for Disease Control and Prevention for providing the expression plasmids of MERS-CoV NPs. We thank Dr. Zheng Fan from Institute of Microbiology Chinese Academy of Sciences for providing the expression plasmids of SARS-CoV NPs.

## REFERENCES

1. National Health Commission of the People’s Republic of China. at http://www.nhc.gov.cn.)

2. Chan-Yeung M, Xu RH. SARS: epidemiology. Respirology 2003;8 Suppl:S9-14.

3. Huang C, Wang Y, Li X, et al. Clinical features of patients infected with 2019 novel coronavirus in Wuhan, China. Lancet 2020;395:497–506.

4. Chan JF, Yuan S, Kok KH, et al. A familial cluster of pneumonia associated with the 2019 novel coronavirus indicating person-to-person transmission: a study of a family cluster. Lancet 2020;395:514–23.

5. Li Q, Guan X, Wu P, et al. Early Transmission Dynamics in Wuhan, China, of Novel Coronavirus-Infected Pneumonia. N Engl J Med 2020.

6. Li X, Zai J, Wang X, Li Y. Potential of large “first generation” human-to-human transmission of 2019-nCoV. J Med Virol 2020.

7. Chen N, Zhou M, Dong X, et al. Epidemiological and clinical characteristics of 99 cases of 2019 novel coronavirus pneumonia in Wuhan, China: a descriptive study. Lancet 2020;395:507–13.

8. Corman VM, Landt O, Kaiser M, et al. Detection of 2019 novel coronavirus (2019-nCoV) by real-time RT-PCR. Euro Surveill 2020;25.

9. Stadler K, Masignani V, Eickmann M, et al. SARS--beginning to understand a new virus. Nat Rev Microbiol 2003;1:209–18.

10. Marra MA, Jones SJ, Astell CR, et al. The Genome sequence of the SARS-associated coronavirus. Science 2003;300:1399–404.

11. Lau SK, Woo PC, Wong BH, et al. Detection of severe acute respiratory syndrome (SARS) coronavirus nucleocapsid protein in sars patients by enzyme-linked immunosorbent assay. Journal of clinical microbiology 2004;42:2884–9.

12. Li X, Mao C, Ma S, et al. Generation of neutralizing monoclonal antibodies against Enterovirus 71 using synthetic peptides. Biochem Biophys Res Commun 2009;390:1126–8.

13. Xiao Q, Wu J, Wang WJ, et al. DKK2 imparts tumor immunity evasion through beta-catenin-independent suppression of cytotoxic immune-cell activation. Nat Med 2018;24:262–70.

14. Li M, An W, Wang L, et al. Production of monoclonal antibodies for measuring Avastin and its biosimilar by Sandwich ELISA. J Immunol Methods 2019;469:42–6.

15. Ma S, Mao Q, Liang Z, et al. Development of a sandwich ELISA for the quantification of enterovirus 71. Cytotechnology 2014;66:413–8.

16. Yu F, Le MQ, Inoue S, et al. Evaluation of inapparent nosocomial severe acute respiratory syndrome coronavirus infection in Vietnam by use of highly specific recombinant truncated nucleocapsid protein-based enzyme-linked immunosorbent assay. Clin Diagn Lab Immunol 2005;12:848–54.

